# RADIANT: A fully configurable radiotherapy dose prediction framework

**DOI:** 10.1101/2025.06.20.25330029

**Authors:** Tucker J. Netherton, Josiane Tchuindjang, Adrian Celaya, Skylar S. Gay, David Fuentes, Laurence E. Court

## Abstract

In this work, we present the Radiotherapy Dose Inference and Analysis Toolkit (RADIANT), an open-source, fully configurable framework for 3D radiotherapy dose prediction. Built upon the Medical Imaging Segmentation Toolkit, RADIANT supports a wide range of network architectures, loss functions, and training strategies. We demonstrate its capabilities using a cervical cancer dataset generated with the Radiation Planning Assistant, consisting of 158 treatment plans. Dose prediction models were trained using five architectures - nnUNet, FMG-Net, W-Net, DDUNet, and Swin UNETR - and evaluated across clinical metrics such as dose score, homogeneity index, and percent errors in D95, D98, and D99. The best-performing model (MAE loss, nnUNet, polynomial scheduler) achieved the lowest average errors across all key dose metrics while also generalizing well to an independent test set. Our results indicate that RADIANT provides a scalable foundation for the rapid development and benchmarking of dose prediction models.

## I. Introduction

**R**ADIOTHERAPY treatment planning is a multi-disciplinary effort that requires medical decision-making, specialized training, and access to imaging and specialized software. Peer review and quality assurance are standard approaches to ensure that each patient receives appropriate, high-quality radiation therapy treatment. In these processes, each treatment plan is reviewed by other radiation oncologists and clinical staff. During this review, medical image-based contours (targets and normal tissues) and three-dimensional (3D) dose distributions are examined.

As demonstrated in multiple clinical trials, poor or non-compliant plans are associated with increased risks of overall mortality and failure of treatment [1], [2]. Although peer review and individualized quality assurance are highly recommended by governing bodies and clinical studies [3]–[5], they are extremely resource-intensive and time-consuming processes.

Recently, deep learning has arisen as a powerful method for predicting patient-specific dose distributions. In the American Association of Physicists in Medicine (AAPM) OpenKBP grand challenge, numerous researchers showcased the ability to accurately predict nominal dose distributions using standard-of-care treatment planning techniques, such as volumetric modulated arc therapy, specifically for head and neck cancer [6], [7]. These predictions have proven feasible for applications in knowledge-based planning and quality assurance, allowing for the detection of suboptimal treatment plans. Consequently, a deep learning-based peer review approach has been proposed. Gronberg et al. [8] illustrated this concept for the head/neck and cervix, demonstrating how comparative dose distributions can facilitate individualized assessments of plan quality. However, one significant limitation is that the quality of dose predictions generated by these supervised deep learning methods is heavily reliant on the training data. Since treatment planning - which includes the contouring of organs and the creation of the plan - remains largely susceptible to manual errors and inter-user variability, aleatoric uncertainty can be considerable, thereby diminishing the utility of predictions. As a result, there is an increasing demand for robust, modular tools that promote reproducible research in dose prediction and enable experimentation with various loss functions, architectures, and data quality control measures.

To address these challenges, we developed the Radiotherapy Dose Inference and Analysis Toolkit (RADIANT), an open-source, fully configurable framework for deep learning–based dose prediction. RADIANT is designed to facilitate reproducible experimentation by providing tools for data preprocessing, model configuration, training, and evaluation across multiple cancer sites and anatomical structures. The framework enables rapid comparison of loss functions, architectures, and optimization strategies while ensuring consistent integration with clinical imaging and contour data. In this work, we demonstrate RADIANT’s capabilities on multilevel dose prediction for cervical cancer, highlighting its potential for generalization, interpretability, and clinical translation.

## II. Previous Work

A number of deep learning models have been developed for radiotherapy dose prediction, with many efforts focusing on head and neck cancer due to the availability of standardized datasets. One prominent example is the 3D Dense Dilated U-Net architecture [9], which was designed for the 2020 AAPM OpenKBP Grand Challenge [6], [7]. This model achieved competitive performance in predicting nominal dose distributions from contours and CT images. However, the architecture and training pipeline were tightly coupled to the OpenKBP dataset and head/neck anatomy, limiting its adaptability to other treatment sites or alternative modeling choices.

Subsequent work by Gronberg et al. extended this approach to individualized quality assurance by comparing predicted dose distributions to clinical plans [8], demonstrating its utility in detecting suboptimal treatment plans. Additionally, a similar strategy was applied by Gronberg et al. to cervical cancer [10], showing that patient-specific predictions could inform plan assessment in anatomically distinct contexts. Despite these promising results, these models were built with fixed architectures, loss functions, and training routines, making it difficult to explore alternative configurations or generalize across disease sites. All of the above studies leveraged the Radiation Planning Assistant (RPA), a web-based, FDA 510(k)-cleared treatment planning system designed to generate high-quality, standardized plans for use in low- and middle-income settings [11]–[14]. While the RPA helped ensure consistency in training data, the models trained on these plans were still subject to dataset-specific design choices and lacked modular implementation.

In parallel, the need for reproducible experimentation in medical image analysis has led to the development of general-purpose frameworks for related tasks like automatic contouring. One such example is the Medical Image Segmentation Toolkit (MIST) [15]–[17], which provides a fully configurable infrastructure for 3D segmentation. MIST enables modular experimentation across model architectures, loss functions, and training strategies, facilitating scalable and reproducible research. While MIST has accelerated development in segmentation, no comparable framework exists for dose prediction - despite similar demands for flexibility, reproducibility, and integration with clinical data.

## III. Methods and Materials

To support reproducible experimentation in dose prediction, we developed RADIANT, a modular framework that enables rapid prototyping and evaluation of deep learning models for dose prediction. In this section, we describe our dataset and RADIANT’s core components, including its preprocessing pipeline, loss functions, model architectures, evaluation metrics, and training configurations.

### A. Patient Data

A total of 158 patients who underwent simulation CT imaging for a gynecological malignancy from 2014 to 2024 were retrospectively analyzed according to a protocol approved by an institutional review board at MD Anderson Cancer Center. Among the patients, 74 had an intact uterus (34 with and 40 without paraaortic node treatment), 53 had undergone hysterectomy (33 with and 20 without paraaortic node treatment), and uterus status was unknown for the remaining 29 (14 with and 15 without paraaortic node treatment).

Using the RPA, cervical cancer treatment plans (using volumetric modulated arc therapy) were automatically generated from each CT, forming a highly consistent dataset of dose distributions, target segmentations, and OAR segmentations. The dose prescription was 45 Gy for the primary PTV and 57.5 Gy for the PTV treating gross nodal disease. Of the 158 datasets, 130 were allocated for training and 28 for an independent test set.

### B. Preprocessing

RADIANT allows the user to input a multidimensional input which includes:

1. 3D imaging (i.e., the treatment planning CT image)
2. Organ-at-risk (OAR) segmentations
3. Planning target volumes (PTV).

For our dataset, OARs included the following: spinal cord, rectum, bladder, femoral heads left and right, kidneys left and right, L4 and L5 vertebral bodies, pelvic bone, sacrum, liver, bowel bag, and body. Targets included the PTV_low_ (receiving 4500 cGy) and PTV_high_ (receiving 5750 cGy).

All input data were interpolated to a standard radiotherapy dose grid size of 2.5 mm × 2.5 mm × 2.5 mm. The OAR segmentations are combined into a single integer-valued array with dimensions *X* × *Y* × *Z* with voxel values ranging from 0 to *n*, where *n* is the number of OARs. Similarly, the PTVs are combined into a single array with dimensions *X* × *Y* × *Z* whose pixel values range from 0 to 1. The PTV voxel values are normalized to have a mximum value equal 1.0 by the maximum dose prescription of 57.50 Gy, and assume there are two dose levels. The preprocessing module in RADIANT then converts all NIfTI files to Numpy arrays for subsequent loading during the training phase.

### C. Loss Functions

There are several loss functions available in RADIANT that capture both voxel-wise accuracy and structure-level dosimetric agreement. First, we implemented the mean absolute error (MAE) and mean squared error (MSE) as voxel-wise losses. These standard regression losses are computed directly between predicted and ground truth dose distributions across all voxels in the volume.

In addition to voxel-level objectives, we incorporated a differentiable dose-volume histogram (DVH) loss that measures agreement between predicted and ground truth DVH curves for each anatomical structure, as proposed in [18]. This loss enables direct optimization of clinical dose metrics and reflects structure-wise coverage. The DVH for a given structure is approximated using a smooth sigmoid-based formulation, making it fully differentiable and compatible with gradient-based learning. The differentiable DVH loss is defined as:

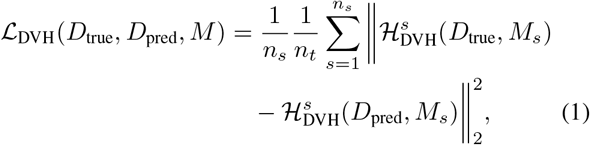

where *D*_true_ and *D*_pred_ are the ground truth and predicted dose distributions, *M*_*s*_ is the binary mask for structure *s, n*_*s*_ is the number of structures, and *n*_*t*_ is the number of dose thresholds. The function 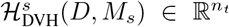 returns the vector of estimated volume fractions receiving at least each threshold dose:

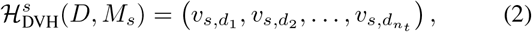

with each term 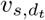 defined as:

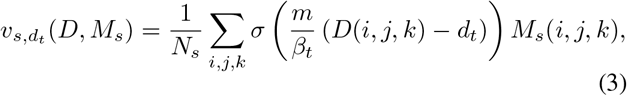

where *σ*(*x*) = 1*/*(1 + *e*^*−x*^) is the sigmoid function, *N*_*s*_ = ∑_*i,j,k*_ *M*_*s*_(*i, j, k*) is the number of voxels in structure *s, m* is a steepness parameter that controls the slope of the sigmoid, and *β*_*t*_ is the dose bin width at threshold *d*_*t*_.

We also incorporated composite loss functions that combine either MAE or MSE with a weighted DVH loss into RADIANT. In this setting, each organ-at-risk (OAR) is assigned an individual non-negative scalar weight *w*_*s*_, allowing greater emphasis to be placed on specific structures. These weighted composite losses encourage both spatially accurate dose predictions and clinically relevant structure-level dosimetric agreement.

### D. Network Architectures

RADIANT includes multiple network architectures that can be selected at runtime, including nnUNet [19], Swin UNETR [20], FMG-Net [16], W-Net [16], and DDU-Net [9]. In addition to these base models, users may optionally enable deep supervision, pocket-size variants for memory efficiency, residual blocks (for CNN-based models), and regularization strategies such as L1, L2, or variational autoencoder regularization.

RADIANT also supports configurable optimization strategies, including Adam and stochastic gradient descent (SGD), as well as a variety of learning rate schedulers. These features are built upon the extensible architecture of the Medical Image Segmentation Toolkit (MIST); for a complete list of supported options, we refer readers to our prior work and public documentation.

Finally, activation functions in RADIANT have been modified to use ReLU in place of softmax, diverging from the original MIST defaults to better suit regression-based dose prediction tasks.

### E. Evaluation Metrics

Following the literature, dose score, mean absolute difference (*MAD*), (*CN*) and homogeneity index (*HI*), *D*_95_, *D*_98_ and *D*_99_ errors were evaluated for each dose prediction. The percent error in each metric (ground truth vs predicted) was also calculated in order to rank all trained models in order of accuracy. These metrics are those typically used in the evaluation of plan quality in radiotherapy.

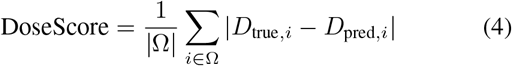

where Ω is the set of voxels in the evaluation region where the dose can be deposited, that is usually the body mask.

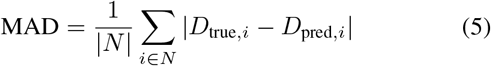

where *D*_*true*_ and *D*_*pred*_ are the true and predicted dose.

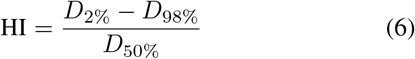

where *D*_*x*%_ is the minimum dose received by *x*% of the target volume.

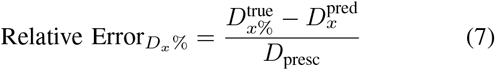

where 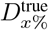 is the ground truth and 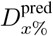 is the predicted minimum dose received by *x*% of the target volume, *D*_presc_ is the prescription for the high target PTV.

## IV. Experiments

### A. RADIANT Training Capabilities

To demonstrate RADIANT’s utility, the following combinations of training options were configured:

- Five neural network architectures: nnUNet, Swin UNETR, FMG-Net, W-Net, and DDU-Net.
- Two loss functions: mean absolute error (MAE) and mean squared error (MSE).
- Four learning rate schedulers: cosine annealing, cosine annealing with warm restarts, exponential decay, and polynomial decay.

All 40 of the resulting models were trained for 1,000 epochs using the Adam optimizer with an initial learning rate equal to 10^*−*3^ and a L2 regularization with the penalty term equal to 10^*−*5^. For the nnUNet, FMG-Net, and W-Net models, we use residual connections and their pocket variants.

### B. Model Refinement

The top 8 performing models were then refined by adding deep supervision (for nnUNet, Swin UNETR, FMG-Net, and W-Net), retraining to 3,000 epochs, and including DVH-weighted loss. Table I shows the top 8 models ranked in ascending order of *D*_95_ errors at the end of training. The complete results for all patients and 40 models are in the supplemental material (supplemental from appendix A). The dose score and *MAD* are metrics that report on the accuracy of the global dose distribution. This implies that overall, the dose predictions from the top models are similar in their 3-dimensional distributions. This is clearly emphasized by 3 with dose scores ranging from 1.32 to 1.5. The dose score for MAE_NNUnet_Polynomial was statistically significant from the ones of other models (*P <* 0.05), except for MAE NNUnet Cosine and MAE_NNUnet_Exponential that both had a slightly higher dose score of 1.38 compared to MAE_NNUnet_Polynomial.

**TABLE I.**
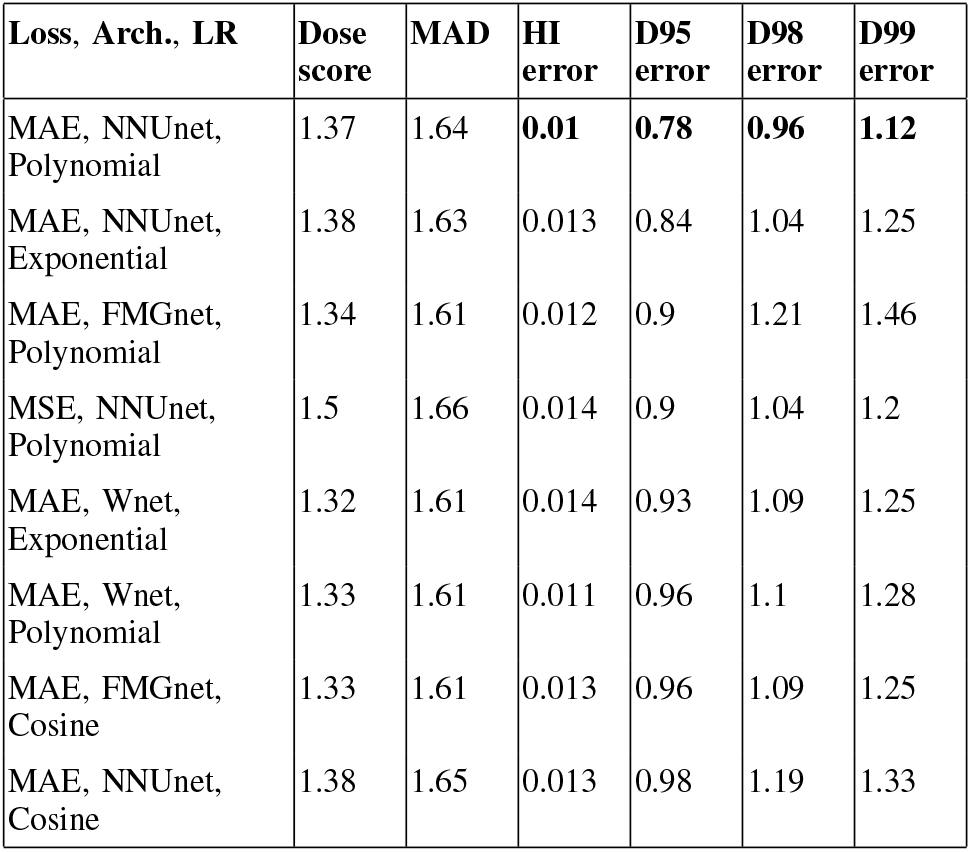
Training and validation experiments. For each model, the average dose score, mean absolute difference (MAD) and homogeneity error, as well as the relative (wrt prescription) errors in ***D***_**95**_, ***D***_**98**_ and ***D***_**99**_ are reported. The lowest error for each metric is in bold. Supplemental material for all 40 models is in the appendix A

**TABLE II.**

| Loss, Model, LR | Dose score | MAD | HI error | D95 error | D98 error | D99 error |
| --- | --- | --- | --- | --- | --- | --- |
| nnU-Net(MAE,poly) | 1.37 | 1.64 | **0.01** | **0.78** | **0.96** | **1.12** |
| MAE, NNUnet, Exponential | 1.38 | 1.63 | 0.013 | 0.84 | 1.04 | 1.25 |
| MAE, FMGnet, Polynomial | 1.34 | 1.61 | 0.012 | 0.9 | 1.21 | 1.46 |
| MSE, NNUnet, Polynomial | 1.5 | 1.66 | 0.014 | 0.9 | 1.04 | 1.2 |
| MAE, Wnet, Exponential | 1.32 | 1.61 | 0.014 | 0.93 | 1.09 | 1.25 |
| MAE, Wnet, Polynomial | 1.33 | 1.61 | 0.011 | 0.96 | 1.1 | 1.28 |
| MAE, FMGnet, Cosine | 1.33 | 1.61 | 0.013 | 0.96 | 1.09 | 1.25 |
| MAE, NNUnet, Cosine | 1.38 | 1.65 | 0.013 | 0.98 | 1.19 | 1.33 |
| MAE, Wnet, CosineWarmRestarts | 1.33 | 1.60 | 0.021 | 1.04 | 1.38 | 1.75 |
| MAE, FMGnet, Exponential | 1.33 | 1.61 | 0.020 | 1.11 | 1.29 | 1.43 |
| MAE, NNUnet, CosineWarmRestarts | 1.38 | 1.65 | 0.020 | 1.14 | 1.49 | 1.82 |
| MAE, FMGnet, CosineWarmRestarts | 1.33 | 1.61 | 0.017 | 1.21 | 1.52 | 1.78 |
| MAE, Wnet, Cosine | **1.31** | 1.60 | 0.015 | 1.29 | 1.53 | 1.79 |
| MSE, Wnet, Exponential | 1.41 | 1.62 | 0.021 | 1.30 | 1.47 | 1.66 |
| MSE, Wnet, CosineWarmRestarts | 1.42 | 1.63 | 0.031 | 1.30 | 1.62 | 1.87 |
| MSE, NNUnet, CosineWarmRestarts | 1.51 | 1.68 | 0.02 | 1.30 | 1.55 | 1.77 |
| MSE, DDUnet, Exponential | 1.35 | **1.58** | 0.021 | 1.36 | 1.75 | 2.04 |
| MSE, NNUnet, Cosine | 1.52 | 1.68 | 0.028 | 1.44 | 1.86 | 2.15 |
| MSE, FMGnet, Cosine | 1.43 | 1.64 | 0.021 | 1.46 | 1.75 | 2.00 |
| MSE, Wnet, Cosine | 1.42 | 1.63 | 0.031 | 1.51 | 1.82 | 2.05 |
| MSE, FMGnet, CosineWarmRestarts | 1.43 | 1.64 | 0.037 | 1.55 | 2.00 | 2.33 |
| MSE, Wnet, Polynomial | 1.41 | 1.63 | 0.035 | 1.56 | 2.00 | 2.34 |
| MSE, FMGnet, Polynomial | 1.44 | 1.63 | 0.028 | 1.70 | 2.08 | 2.41 |
| MSE, NNUnet, Exponential | 1.47 | 1.68 | 0.024 | 1.73 | 2.18 | 2.57 |
| MSE, DDUnet, Cosine | 1.35 | 1.60 | 0.026 | 1.74 | 2.16 | 2.58 |
| MSE, FMGnet, Exponential | 1.43 | 1.64 | 0.028 | 1.79 | 2.07 | 2.34 |
| MAE, Unetr, Exponential | 1.36 | 1.64 | 0.034 | 1.96 | 2.69 | 3.29 |
| MAE, Unetr, Polynomial | 1.39 | 1.67 | 0.034 | 1.99 | 3.07 | 3.84 |
| MAE, DDUnet, Cosine | **1.31** | 1.60 | 0.035 | 2.02 | 2.61 | 3.07 |
| MAE, DDUnet, Polynomial | **1.31** | 1.60 | 0.038 | 2.10 | 2.73 | 3.11 |
| MAE, DDUnet, Exponential | **1.31** | 1.61 | 0.034 | 2.22 | 2.96 | 3.48 |
| MAE, Unetr, Cosine | 1.38 | 1.65 | 0.034 | 2.30 | 3.10 | 3.64 |
| MSE, Unetr, Cosine | 1.47 | 1.67 | 0.052 | 2.36 | 3.08 | 3.64 |
| MSE, Unetr, Polynomial | 1.46 | 1.67 | 0.036 | 2.57 | 3.05 | 3.50 |
| MSE, DDUnet, Polynomial | 1.37 | 1.61 | 0.031 | 2.66 | 3.10 | 3.48 |
| MAE, Unetr, CosineWarmRestarts | 1.36 | 1.65 | 0.054 | 2.67 | 3.72 | 4.48 |
| MAE, DDUnet, CosineWarmRestarts | 1.32 | 1.60 | 0.044 | 2.79 | 3.56 | 4.05 |
| MSE, Unetr, CosineWarmRestarts | 1.47 | 1.68 | 0.051 | 2.96 | 3.85 | 4.43 |
| MSE, DDUnet, CosineWarmRestarts | 1.36 | 1.61 | 0.043 | 3.28 | 3.98 | 4.42 |
| MSE, Unetr, Exponential | 1.48 | 1.67 | 0.035 | 3.50 | 4.08 | 4.54 |

The doses to select PTVs and OARs represent localized portions of this dose distribution. The average dose error across all 8 models was within 1% for PTVs and 2.5% for OARs (Figure 4)-indicating high accuracy at many relevant focal areas. This is further emphasized by Figure 2 in the predicted versus ground truth DVH curves of a randomly chosen example patient. The variation in average maximum dose across all 8 models was slightly larger (Figure 1). This is expected as deep learning base dose prediction is known to generalize and smooth predictions relative to ground truth data, making prediction of global maxima imprecise. Because PTV dose coverage (i.e. *D*_95_, *D*_98_) is an important metric of plan quality, the top models were ranked by *D*_95_ error (Table I). This is analogous to the accuracy in the “shoulder” portion of the DVH curve for the PTV.

**Fig. 1.**
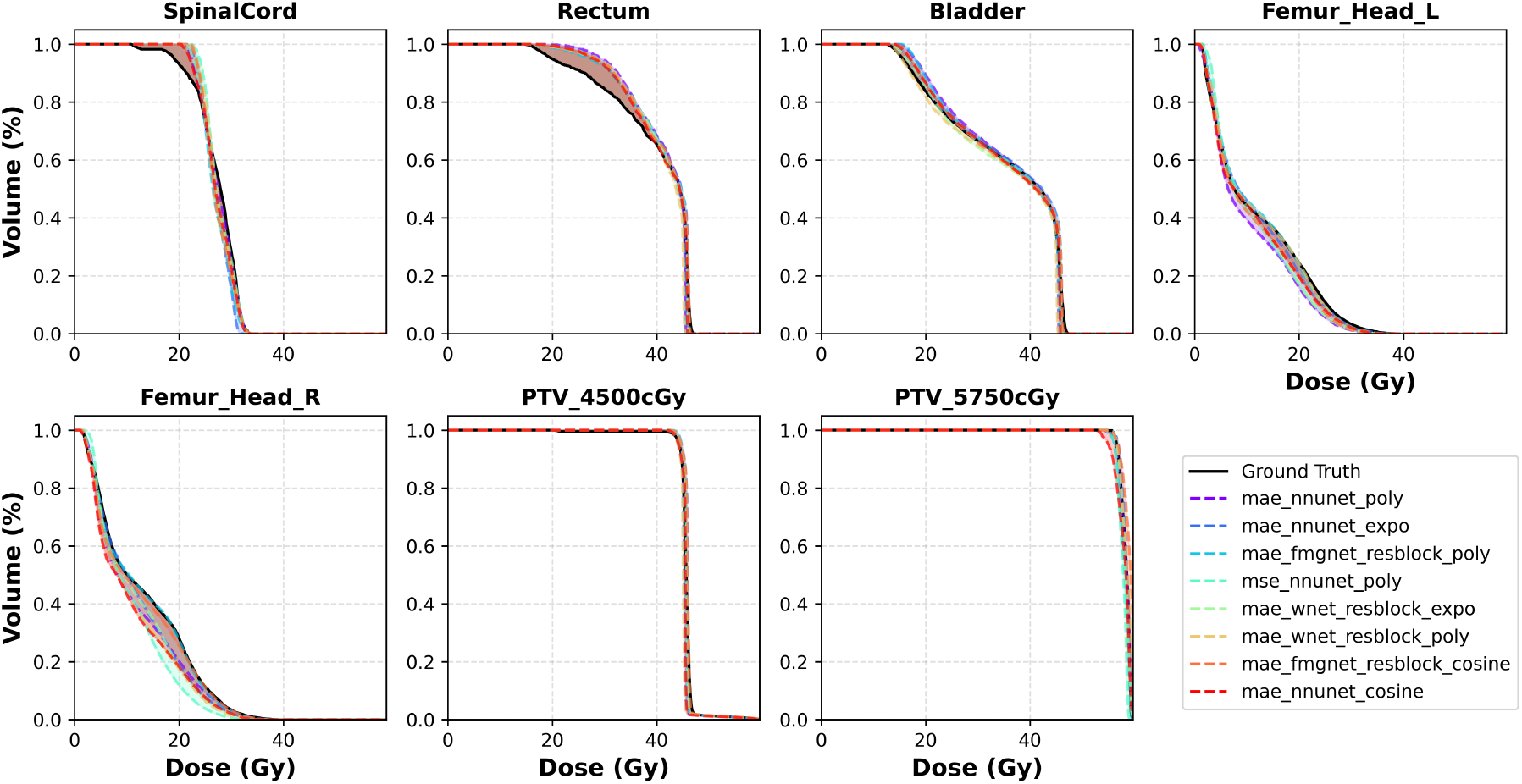
Dose-volume histograms (DVH) curves for some key OARs and both targets for all models. Ground truth DVH is in solid black, predictions are in dashed colors.

**Fig. 2.**
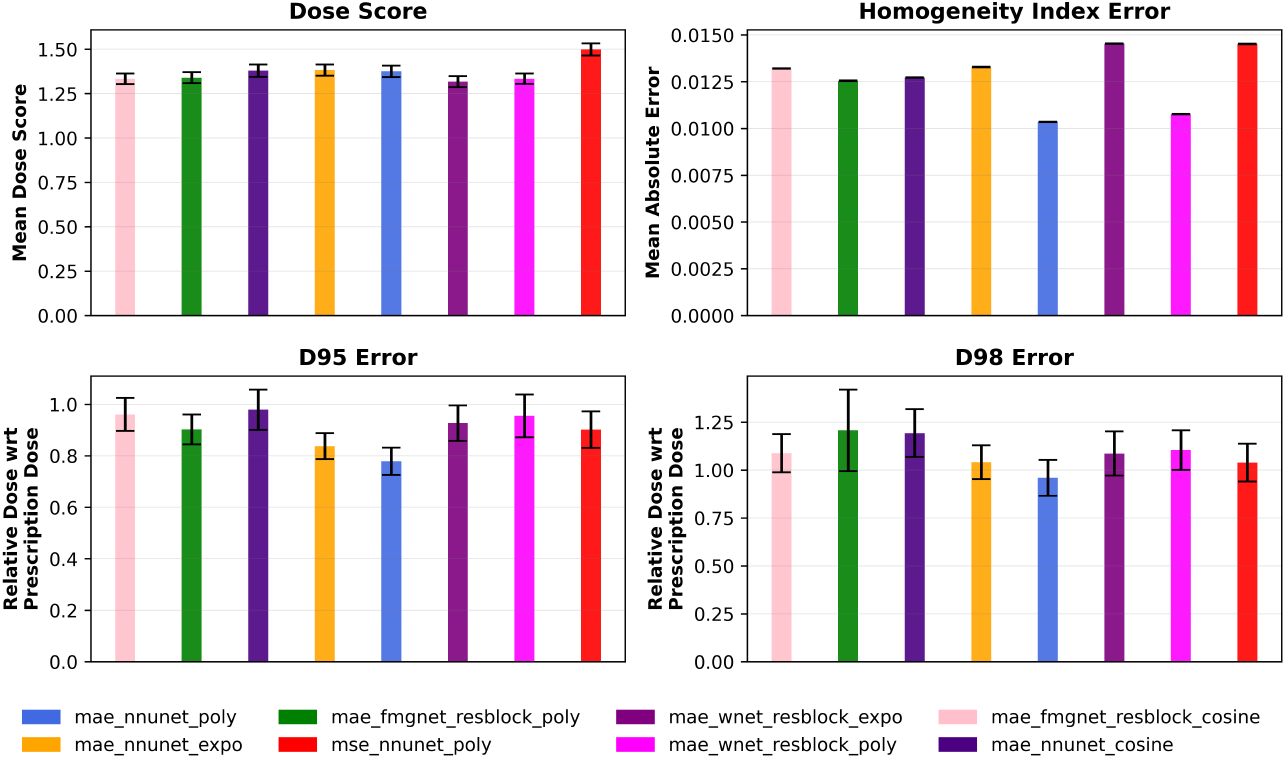
Mean average absolute error for for dose score, homogeneity index. Relative dose error wrt percent of prescription dose for ***D***_**95**_ and ***D***_**98**_. The error bars represent the 99% confidence interval for all training patients for each model.

**Fig. 3.**
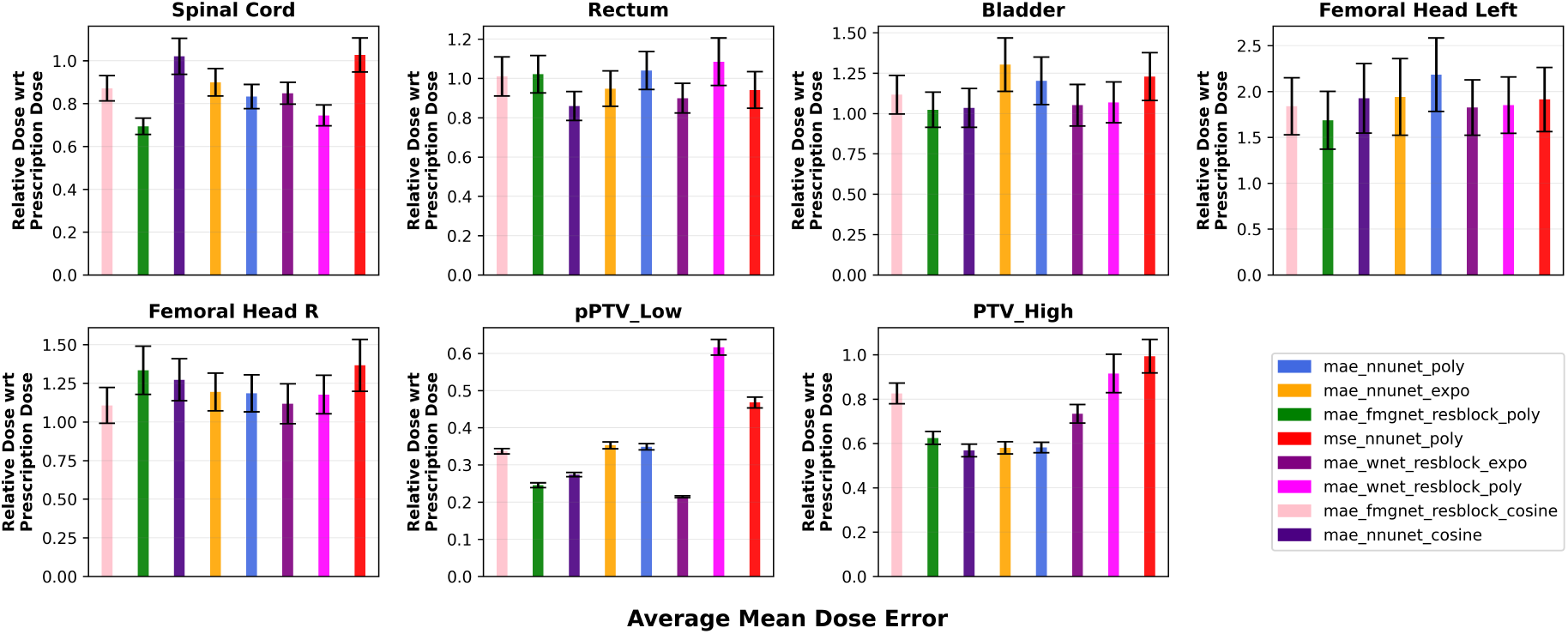
Prediction errors in mean dose for both targets and some key OARs. The error bars represent the 99% confidence interval for all training patients for each model.

**Fig. 4.**
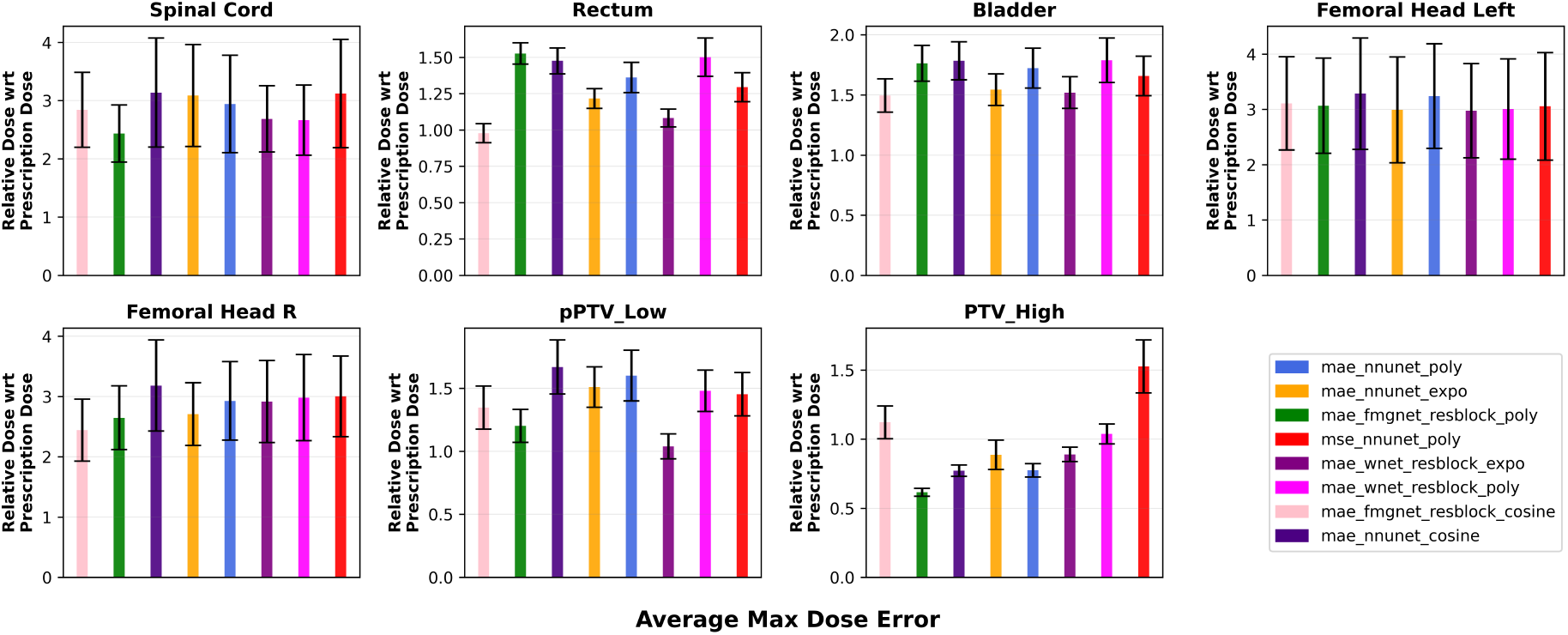
Prediction errors in max dose for both targets and some key OARs. The error bars represent the 99% confidence interval for all training patients for each model. ***D***_***max***_ is the max dose within 0.2 cm^**3**^ volume.

The nnU-Net model trained with MAE loss and polynomial LR scheduler had the lowest errors in predictions of *HI* (0.01 ± 0.009%), *D*_95_ (0.78% ± 0.66%), *D*_98_ (0.96% ± 0.92%) and *D*_99_ (1.12% ± 1.11%). The training and loss curves for this final model are shown in Figure 10. In addition to this top model (nnU-Net, MAE, poly), this same configuration was retrained with the MAE + wDVH loss and was also used in the test set DVH analysis in the proceeding section.

### C. Dose-Volume-Histogram Analysis

Figure 5a shows the ground truth and predicted dose for a patient with the para-aortic nodes being treated and boosted to 5750 cGy (top), the pelvis was treated to 4500 cGy (bottom).

**Fig. 5.**
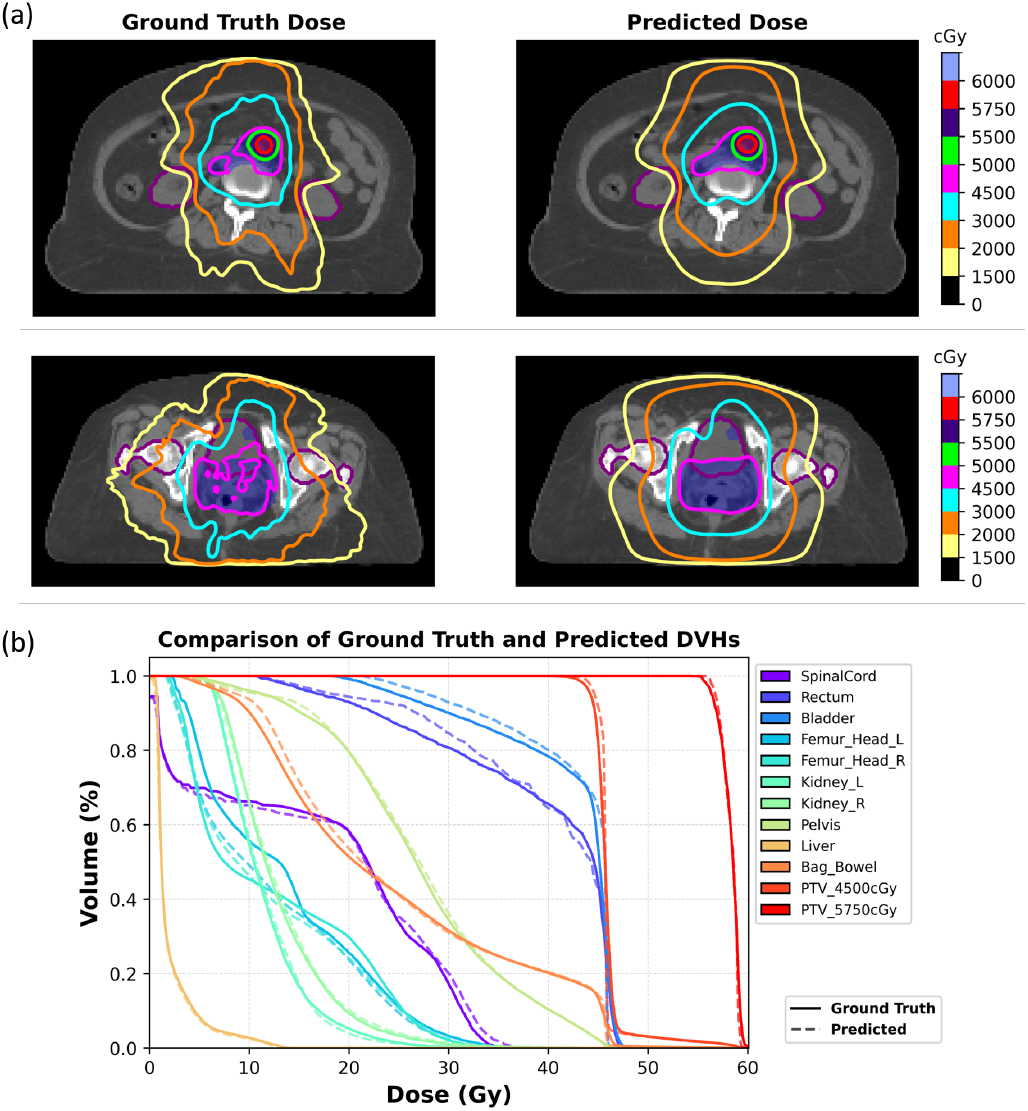
Comparison between ground truth RPA data and predicted dose distributions for a test set patient using model MAE_NNUnet_Polynomial. a) Top: Dose distributions in the PAN are shown with the bladder and left and right kidney organs (purple contours). The boost in PTV_5750cGy is shown in red and the other target PTV_4500cGy is plotted in blue. a) Bottom: Pelvis area for same patient has no boost, only dose of 4500 cGy to the low target. Bladder and left and right femoral heads are plotted in purple. Countour lines are isodose lines. b) Comparison of DVH curves between RPA ground truth (solid line) and predicted dose (dashed line). OARs and both targets are plotted.

The DVH was plotted for the same patient in Figure 5b with some key OARs. Plots a and b of Figure 6 show the dose and DVH predictions for a patient who only had the cervix treated to 4500 cGy and pelvic nodes boosted to 5750 cGy. This shows that the model is able to perform well independent of where the boosted nodes are in the patient. For both figures 5a and 6a, the high dose target PTV_5750cGy is contoured in red, the low dose target PTV_4500cGy is contoured in blue. For the PAN area (Figure 5a top), the left and right kidneys are contoured in purple. For the pelvis area (Figure 5a (bottom) and Figure 6a), the bladder and both femoral heads are contoured in purple. The DVHs for all structures show that the predictions (dashed lines) closely follow the RPA ground truth (solid lines), with a great match in the two targets as well as OARs.

**Fig. 6.**
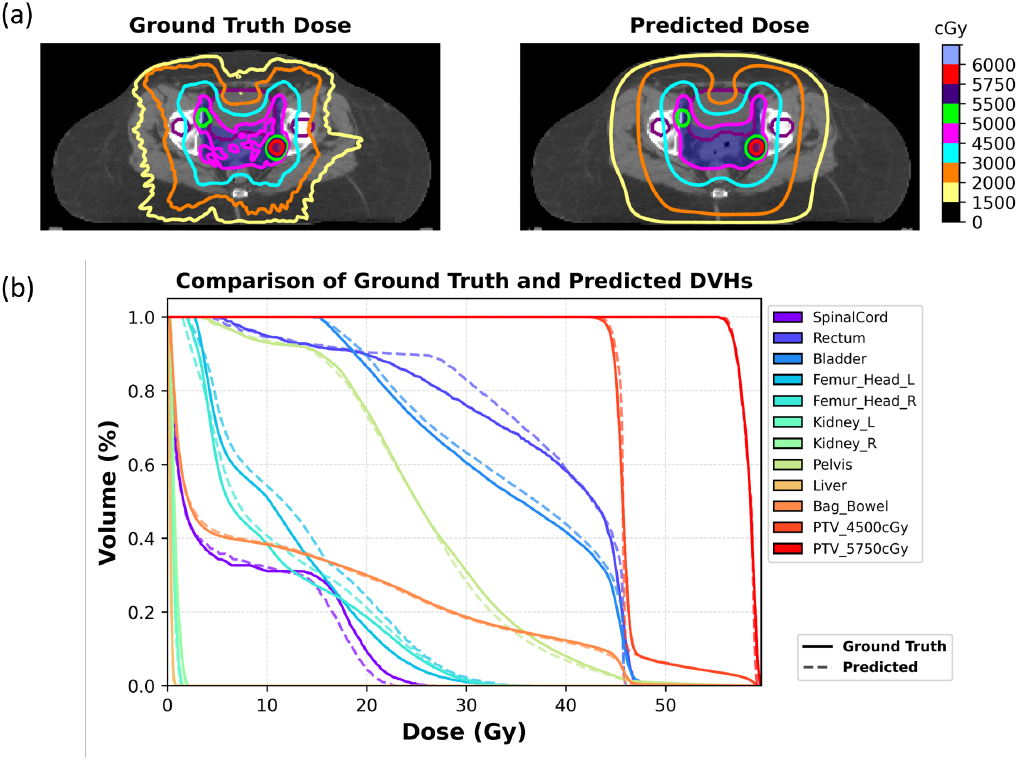
Comparison between ground truth RPA data and predicted dose distributions for a test set patient using model MAE_NNUnet_Polynomial. a) Dose distributions in Pelvis area for another patient with boost, only dose of 4500 cGy to the low target. Bladder and left and right femoral heads are plotted in purple. Countour lines are isodose lines. b) Comparison of DVH curves between ground truth (solid line) and predicted dose (dashed line). OARs and both targets are plotted.

We also plotted for the same patients the results of the model MAE_wDVH_NNUnet_Polynomial that was run for 3000 epochs. Adding the weighted DVH to the top performing model MAE_NNUnet_Polynomial also showed similar quality of results with it as we can see in Figures 7 and 8.

**Fig. 7.**
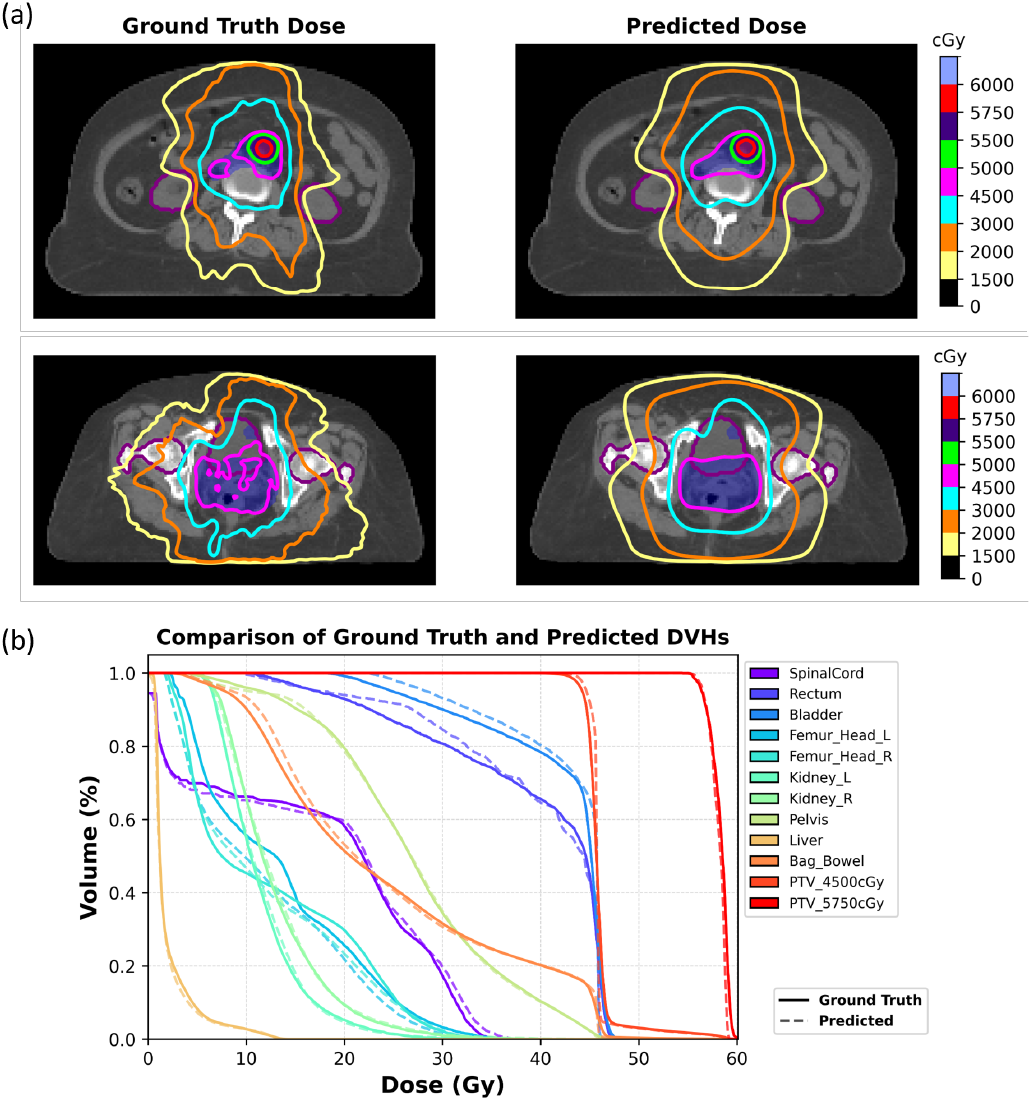
Comparison between ground truth RPA data and predicted dose distributions for a test set patient using model MAE_wDVH_NNUnet_Polynomial. a) Top: Dose distributions in the PAN are shown with the bladder and left and right kidney organs (purple contours). The boost in PTV_5750cGy is shown in red and the other target PTV_4500cGy is plotted in blue. a) Bottom: Pelvis area for same patient has no boost, only dose of 4500 cGy to the low target. Bladder and left and right femoral heads are plotted in purple. Countour lines are isodose lines. b) Comparison of DVH curves between RPA ground truth (solid line) and predicted dose (dashed line). OARs and both targets are plotted.

**Fig. 8.**
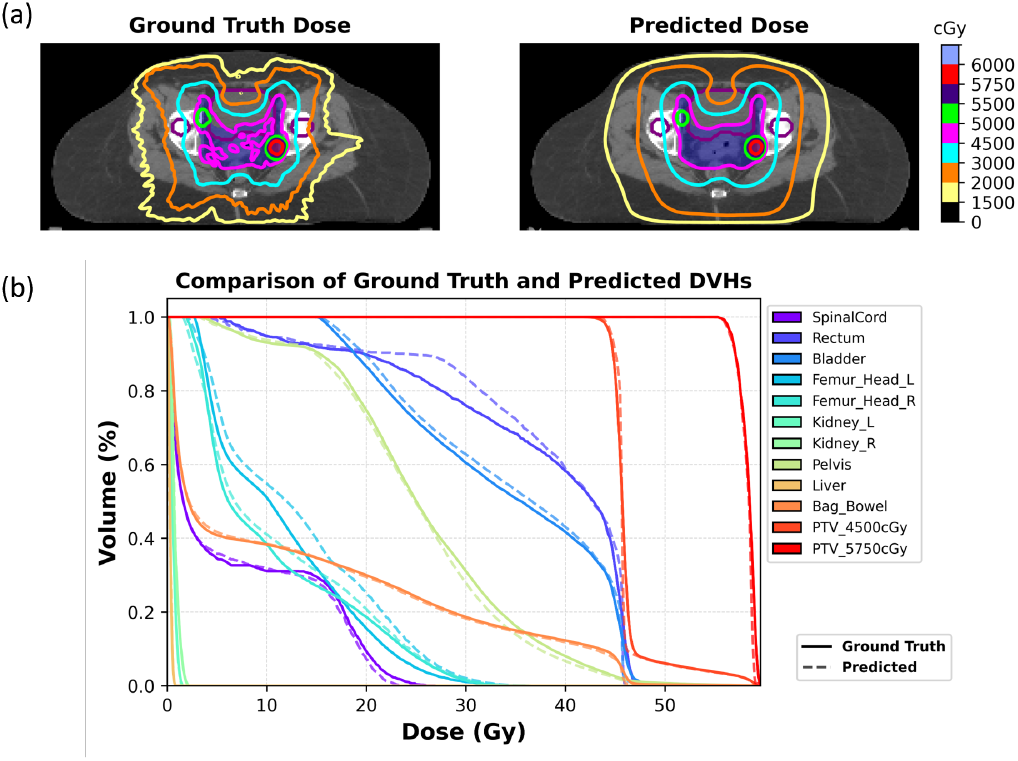
Comparison between ground truth RPA data and predicted dose distributions for a test set patient using model MAE_wDVH_NNUnet_Polynomial. a) Dose distributions in Pelvis area for another patient with boost, only dose of 4500 cGy to the low target. Bladder and left and right femoral heads are plotted in purple. Countour lines are isodose lines. b) Comparison of DVH curves between ground truth (solid line) and predicted dose (dashed line). OARs and both targets are plotted.

In Figure 9 we plot the DVH curves for a patient for whom the model performed poorly. The left plot is the DVH curve for the model MAE_NNUnet_Polynomial and the right plot is the DVH curve for the model MAE_wDVH_NNUnet_Polynomial. For both plots, the predicted DVH (dashed lines) misses the ground truth DVH (solid lines) for the high dose target. Adding the DVH loss did not improve the results for the target, though it did for the OARs.

**Fig. 9.**
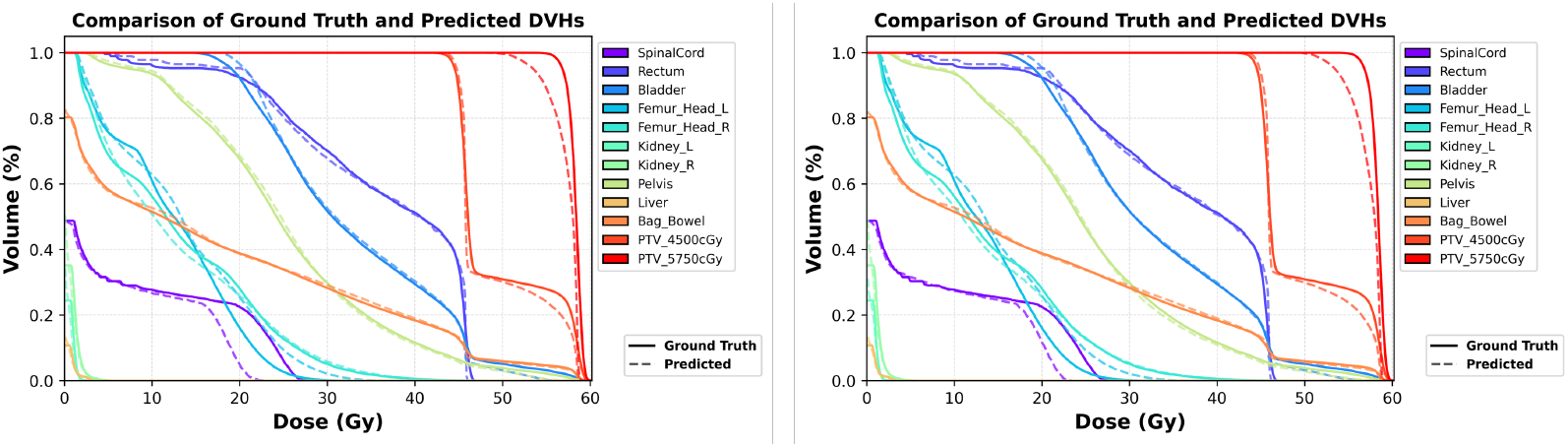
Comparison of DVH curves between ground truth (solid line) and predicted dose (dashed line) for a test set patient where both models MAE_NNUnet_Polynomial (left plot) and MAE_wDVH_NNUnet_Polynomial (right plot) performed poorly on the high dose target.

**Fig. 10.**
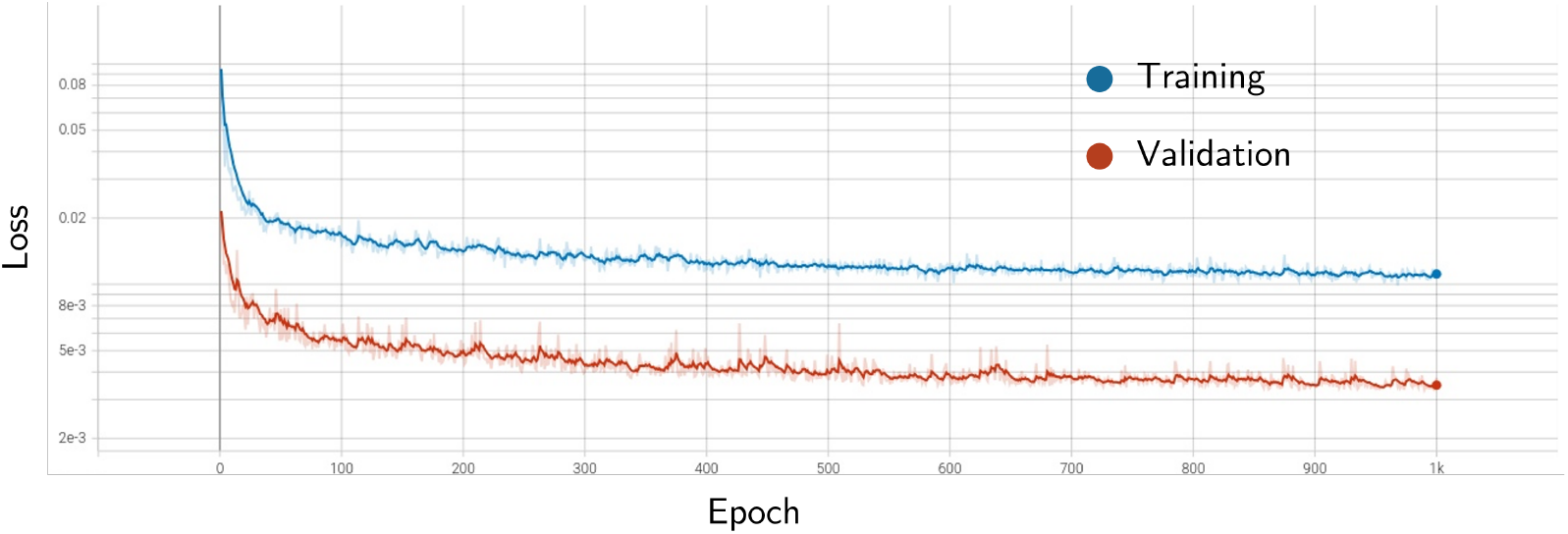
Training and validation loss curves for the top performing model (nnUNet, MAE, Polynomial). Here we see that that the validation loss curve is consistently lower than the training loss curve. This indicates that our best model is able to generalize well to unseen data. We see similar curves for the other top performing models.

## V. Discussion and Conclusions

In this study, we introduced RADIANT, a modular and extensible framework for 3D radiotherapy dose prediction, and demonstrated its utility through a comprehensive evaluation on a large, high-quality cervical cancer dataset. Our results show that RADIANT enables rapid experimentation across a wide range of model architectures, loss functions, and training strategies, while maintaining high accuracy in clinically relevant dose metrics.

Among the 40 trained models, the nnU-Net architecture with MAE loss and polynomial learning rate scheduler consistently outperformed others across global and structure-specific metrics, achieving sub-1% errors in *D*_95_ and *D*_98_ and a homogeneity index error of 0.01. These results highlight the effectiveness of combining robust architectures with carefully selected optimization strategies for dose prediction tasks. Furthermore, the incorporation of *DVH-weighted loss* in the refinement phase improved structure-level agreement, particularly for organs-at-risk (OARs), without compromising global dose accuracy.

The high consistency of dose predictions across the top performing models, as shown by the low dose score and MAD values, suggests that RADIANT can produce clinically meaningful dose distributions with minimal deviation from the truth of the ground. In particular, the mean dose error was within 1% for PTVs and 2.5% for OARs. However, as expected, prediction of global maxima showed greater variability, reflecting the known tendency of deep learning models to generalize and smooth isodose lines when compared to doses from individually optimized treatment plans. Furthermore, the top models studied in this work use the “pocket” implementation [15], which uses substantially less parameters (700k in total for pocket nnU-Net) than those in the original nnU-Net implementation. That the final model generalizes well while operating on less GPU memory is a strength of RADIANT which can be further leveraged by other researchers and developers using a variety of hardware.

Our findings also highlight the importance of model selection and training configuration in dose prediction. While all five architectures were capable of learning meaningful dose distributions, pocket nnU-Net consistently demonstrated superior performance, likely due to its dynamic adaptation to dataset-specific properties and its proven success in medical image segmentation tasks. The use of composite loss functions that integrate voxel-wise and structure-level objectives further enhanced clinical relevance, allowing for targeted optimization of critical anatomical regions.

By providing a unified platform for dose prediction, RADIANT addresses a key gap in the field: the lack of reproducible, configurable, and scalable tools for benchmarking and deploying deep learning models in radiotherapy (Github avialble upon article acceptance).

### A. Limitations and Future Work

While RADIANT demonstrates strong performance on a well-curated cervical cancer dataset, further validation on multi-institutional and multi-site datasets is necessary to assess generalizability. Additionally, future work will explore uncertainty quantification, physician review, multi-modal inputs (e.g., MRI, PET), and integration with clinical decision support systems through the RPA.

## Data Availability

Datasets are not available, but github code will be publicly available upon article acceptance

https://github.com/mist-medical/MIST

https://researchrpa.mdanderson.org/

## Appendix and the Use of Supplemental Files

### A. Table of results of all 40 models

## Acknowledgment

The authors would like to acknowledge the MD Anderson Enterprise AI team, the Radiation Planning Assistant team, MD Anderson Global Oncology Program, and the MD Anderson Institute of Data Science in Oncology.

## Notes

This work was submitted on XX/XX/XXXX. “This work was supported in part by”. TKN acknowledges support of the National Institute of Health Loan Repayment Award. SSG received support from the Cancer Answers Scholarship, the Larry Deaven PhD Fellowship, and the Dr. John J. Kopchick Fellowship.

### Competing Interest Statement

The authors have declared no competing interest.

### Funding Statement

This study was funded in part by a Cancer Center Support Grant (CCSG) from the National Institute of Health, National Cancer Institute under award number P30CA016672.

### Author Declarations

These data were retrospectively analyzed according to a protocol approved by an institutional review board at MD Anderson Cancer Center. Consent was waived due to the retrospective nature of the study.

